# Pandemic, Primary Health Workers and work trajectories: A Memoing Representational Study

**DOI:** 10.1101/2021.11.19.21264459

**Authors:** K Rajasekharan Nayar, Ujjwala Gupta, Bindhya Vijayan, Sunitha SB, Kapila VS, Shringa Sivanand, Krishna SH, Shaffi Fazaludeen Koya

## Abstract

More than 65 per cent of the population in India lives in rural areas with the highest overall burden of disease. The Indian rural health care system is composed of the three-tier system comprising Sub-Centres, Primary Health Centres, and Community Health Centres with a considerable shortfall in health facilities at different levels - 18 per cent at Sub centre level, 22 per cent at PHC level and 30 per cent at CHC level. The real facts and figures of the epidemic in rural areas are not known yet except for broad distribution patterns. The course of events with respect to preventive strategies to control COVID-19 especially the experiences in states like Kerala which has comparatively well-developed health infrastructure are important as a lesson for managing future emergencies. In the present study, the responses and experiences of the frontline health workers including ASHA workers toward the pandemic are documented. We followed a Memoing approach- largely similar to in-depth interviews- based on conversations with primary level health workers including ASHA workers and Junior Health Staff. The conversations which lasted for about one hour and in some cases more were presented by the respondents as experiential representations and memoranda on which reflective notes were prepared by the authors who conversed with the staff. The conversations mainly echoed the complaints, concerns and criticisms of the staff regarding the program and the severe limitations that they faced in COVID control. Based on the narratives as well as representations, we could identify interlinked dominant and minor specific-context related issues which are important for equity-based universal health coverage. Firstly, training of primary health workers in Primary Emergency Health care is important in order to counter unpleasant human interactions and also for maintaining security. The training is also important to counter misinformation which is hampering positive health actions. Secondly, it is important to reinvigorate the medical loop and preventive protocols in health programs to strengthen the health service system at the grassroots level especially enhance the trust between the workers and the people. COVID 19 pandemic is an opportunity to recognize and reinforce the role of primary care workers and formulate gender-sensitive and effective control strategies.

---

More than 65 percent of India’s population lives in rural areas with the highest overall burden of disease. The Indian rural health care system is composed of three-tier system comprising Sub-Centres, Primary Health Centres, and Community Health Centres with considerable shortfall in health facilities at different levels - 18 percent at Sub centre level, 22 percent at PHC level and 30 percent at CHC level (as on March 2018)^1^. Workforce shortage is also substantially high despite expansion of infrastructure. Rural India has 3.2 government hospital beds per 10,000 people. Many states have a significantly lower number of rural beds than the national average. The state of Uttar Pradesh has 2.5 beds per 10,000 people in rural areas. Rajasthan and Jharkhand are at 2.4 and 2.3, respectively. Maharashtra, which had witnessed the largest number of cases, has 2 beds while Bihar has only 0.6 beds ^2^. It is against such a scenario that the participation of community health care workers to tackle public health emergencies such as COVID -2019 has to be assessed. The rural health care system is not adequate and prepared to contain COVID-19 transmission in the rural areas, especially in many North Indian States because of the shortage of doctors, hospital beds, equipment etc. It is a wakeup call and what is important at this moment is to use the lessons of this pandemic to improve the health care systems in rural areas considering the huge population and untrained staff in caring and handling of patients during an outbreak of infectious diseases.

The real facts and figures of the epidemic in rural areas are not known yet except broad distribution patterns. The course of events with respect to preventive strategies to control COVID-19 especially the experiences in states like Kerala which has comparatively well-developed health infrastructure are important as a lesson for managing future emergencies. Kerala with historic and internationally acclaimed achievements in the health sector have become a victim of this transition. Despite the positive image of the health and social sectors and considerable investments in the health sector and establishment of modern care institutions, Kerala a has complex epidemiological scenario largely due to the emergence of new diseases such as seasonal fevers, chikungunya, Dengue, and other viral infections in epidemic proportions especially during and after the monsoon season^3^. Changing life styles of the population, the new food culture and the large migratory population in the state have also played a significant role in the new epidemiological profile of the state. This new epidemiological scenario exists in the backdrop of all-round commercialization of health care and de-emphasizing public health and primary care^4^. All these factors need to be understood through empirical studies which only could provide an evidence-based public health policy framework as against an eminence-based decision making which is rampant with regard to public health programs

The health service is a socio-technical institution and there exists a number of actors and a multitude of interests which influence the decision-making and functioning within such an institution^5^. A techno-centric approach to public health is dominant universally and nationally. But there also exits an alternate vision to public health which considers health services as a social and cultural institution and which gives primacy to primary health care. Recognising the role of Community Level Workers form part of this vision and it is during the pandemic that the importance of such workers has been felt both by the civil society and the government. Workers such as ASHA and other frontline health workers have a great role and responsibility in the pandemic control apart from maintaining the routine health activities. In many countries, they have been used mainly for community preparedness, health maintenance and educational activities. It is now known that many such workers have faced great personal risks without adequate personal protective equipment to combat the pandemic^5^. In a country which is facing severe shortage of health manpower, this is a serious matter of concern. It is important to understand how they performed their roles and how the community received it in order to devise suitable emergency governance in future.

The state of Kerala in India had a favourable history of maintaining better health of the population as well as in containing sporadic small episodes of new illnesses. Initially, the availability of a committed medical force certainly helped in containing the virus to some extent. The role of primary care workers in control activities is not small and it is important to document their experiences and work trajectories especially from this state which saw the first arrival of COVID-19 case.

## Methodology

In the present study, the responses and experiences of the frontline health workers including ASHA workers toward the pandemic are documented. We followed a Memoing approach- largely similar to in-depth interviews- based on conversations with primary level health workers including ASHA workers and Junior Health Staff. The conversations which lasted for about one hour and in some cases more were presented by the respondents as experiential representations and memoranda on which reflective notes were prepared by the authors who conversed with the staff. The conversations mainly echoed the complaints, concerns and criticisms of the staff regarding the program and the severe limitations that they faced in COVID control. These were categorised and then abstracted to represent the worldview of the primary health care workers fully engaged in the difficult and intensive control activities. For this purpose, we selected five different districts from the state of Kerala, namely, Thiruvananthapuram from the southern part, Alappuzha and Kochi from the mid-region and Kozhikode and Malappuram from the North. Most of these districts witnessed considerable surge in cases. Overall, we could cover 41 primary health care workers despite their busy schedule. In order to understand an entirely contrasting scenario, we also interacted with a few frontline workers from the state of Jharkhand in Northern India from both urban and rural areas which also witnessed a surge in cases when large number of migrant workers started returning to the state^6^. This also helped in assessing contextual validity of the qualitative tools.

**Figure 1.**
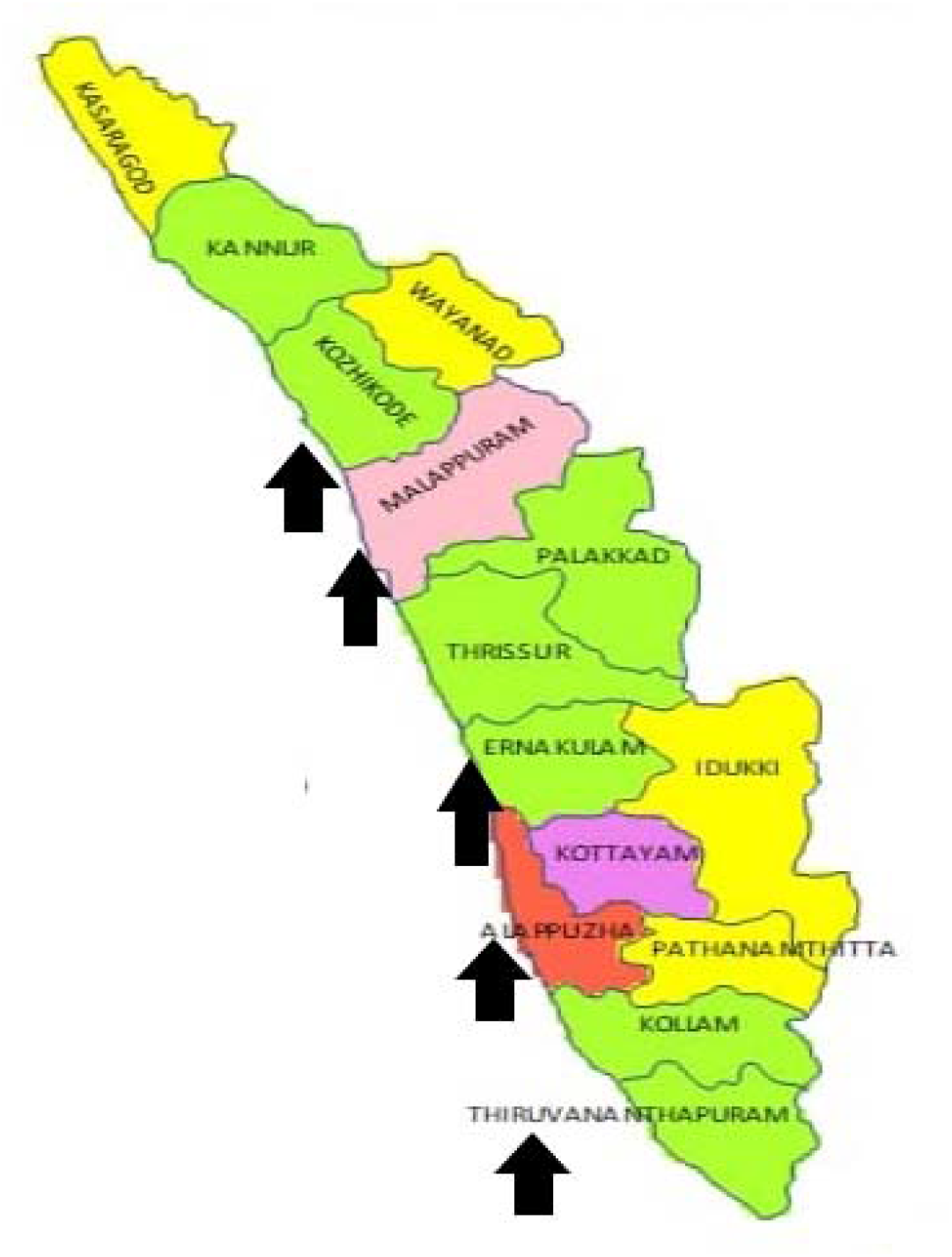
Map Kerala and the districts covered

## Work Trajectories and Representations in different districts of Kerala

### Issues regarding Fieldwork

Most frontline workers mentioned that fieldwork is the heavy work and felt really cumbersome during the pandemic period. Contact tracing is another tedious issue as people are not willing to share information in many instances. Workers revealed that they were involved in the preventive activities almost most of their working time. Many workers revealed that normal responses from the people were lacking. Social isolation was one of the important phenomena felt by most workers as they were not allowed to enter many houses for classes and instructions.

*“People also interacted with us differently as everyone talked to us from a distance. But we managed this isolation even within our own houses”*

Fieldwork by the health workers is an important dimension of pandemic control activities. According to a Health Inspector, ASHA worker performed to the fullest potential during this period especially to detect positive people and to provide all the help to them. But these activities according to health workers were not very smooth.

Despite high health literacy some places in the state also faced resistance to home visits. *“People sometimes expressed displeasure when we used to regularly visit the homes to enquire about their health. However, we used to handle the situation somehow. House visits are one of the most tedious tasks”*.

One ASHA worker said, *‘It was difficult to do fieldwork during this period. But I completely involved myself in home visits as much as I could although personally I have difficulties. I also have a family and a small child. Therefore, I also need to be careful and take protections myself”*

Another worker, said, “*In some areas, I faced bad behavior when I went to enquire about a COVID case. As women workers we face such behavior more often. The people insulted me and said I am spreading the disease as a health person*”. Many workers also mentioned lack of compliance with their instructions especially regarding maintaining social distance or even with respect to testing. Workers revealed that this is largely due to misinformation that people receive through the social media. This trend is common in all the districts that were covered.

It is important to realize that complete commitment of these primary care workers helped in sustaining control activities and ensuring that people would comply with the necessary preventive actions needed from them.

### Misinformation

Misinformation regarding the infection through the social media was certainly an important problem which negatively influenced the preventive work including vaccination. According to one ASHA worker, *“When we get the information regarding such cases, the local NGOs and other organizations are contacted and the issue is sorted out to some extent”*.

According to one Junior Public Health Nurse, *“it is easy to resolve misinformation if we have strong connections (including ASHA and other primary workers) in the community and we always inform them about those persons. We have formed a ‘COVID awareness’ committee for that purpose. The Panchayat Member is the chairman of the committee”*.

Another worker informed, *” We used to handle this by giving correct information by house visits and vehicle announcements about the infection. Giving correct information and awareness to the people was always a challenge as information available to us was not complete”*.

One health worker said, *“we stand near the gate or entrance and give the instructions”*

The influence of social media is so strong in the state as for all information people depend on it and people used to spread the information by forwarding all the messages they receive to other people.

### Work pressures and additional duties

Pressure to complete the existing tasks and the additional duties due to pandemic control is a serious problem which affected the women workers. This has affected many people mentally and physically and some of them have been finding it difficult to carry out their work. The dual role people had to perform in terms added work pressure and demands from the family had also significantly impacted their work trajectory.

One Junior Public Health Nurse said that routine activities like NCD clinics have been affected. “*We need to complete the targets in according to the time. Now we are actually doing the double work after the COVID -19. I do the paper work after mid night”*.

One health worker said, “*although we have this much work load there is no increment or added benefits for us. Even today I do all these without pending since this is my passion. I love to do community services. Those smiling faces give me the completeness”*.

*“When we go and ask people who are infected to go on quarantine, they ask us who we are to ask them. That is the situation now. We do all the work and we face all the bad mouth. Even there are some in the non-infected who do not listen to us”*.

*“They tell us that it is difficult to follow this life pattern always. Some people shouted at us but the police helped us to maintain the protocol*

Some workers also faced unpleasant situations during their work. One ASHA worker narrated her experience-*“I was worried and sad when a young boy had beaten me for telling him to wear the mask. My family got tensed about my job due to this experience. The incident also made me depressed. In another incident, I faced violence and a group of guys had called abusive words. This was a major problem during the pandemic period but most of the ward representatives stood by me”*.

Personal trajectories also matter to many community level workers as many of them are young, married and have small children. *“My role in community is tough, but I am supposed to do my work, have to deliver medicines and give proper information. I am worried too about my family as I am always exposed to all kind of infection. Phone calls at midnight and sleepless nights were so common during COVID-19 lockdown period. That was also a problem I had faced during COVID time”*.

## The case of Jharkhand

Jharkhand which is predominantly a tribal state, with poor socio-economic development was hit severely by the pandemic like other states of India. The state witnessed fatal second wave of COVID – 19 that put additional pressure on previously ill-equipped health infrastructure to the extent that it increased the deprivation of medical care even for critical patients 8. With the ability of ASHA (named as Sahiya in the state) to access and reach women in the community, they were burdened with the task of surveillance of COVID active cases, tracking migrants and to improve coverage. Some of the problems such as work pressures and negative responses of the people observed in the state were similar to the pattern in Kerala. We present some of the responses of the ASHA workers who were engaged in detailed conversations with us.

### Non-COVID duties

Added to the high number of COVID cases of, Jharkhand is also prone to vector borne and water borne diseases like dengue, malaria and typhoid that made the situations worse for the health care system to handle. One Sahiya says-

“Last time, we were used for survey, testing and community mobilization, and in this phase we are involved in COVID vaccination drive and other additional duties related to dengue and malaria which are also on the rise.”

### Issues of safety in work and new work pressure

During the second wave, situation in rural Jharkhand remained even more debilitating, with no definite estimate of the upsurge in cases and mortality rate. Sahiyas complained of poor quality protective equipment being used especially for them.

“Last time we were given some masks and gloves which got old and torn off. Now, most of us are either buying our own or using thin clothes like our stole, duptatta as mask which are neither safe for ourselves nor anyone else. We actually feel left out and very inferior in the system where most of the privileges are given to doctors and nurses in full time employment.”

“In the first phase of COVID, we did not get any training but our role remained instrumental in testing, mobilizing and arranging for setting up quarantine centers since the active cases were less but with the arrival of second wave most families are completely infected and that frightens us to go inside their houses and ask. We only knock their doors from outside and ask, no matter how far correct information

“several Sahiya workers got infected while working, a few died. But our loss while working in this disaster is neither in any counts nor it matters to anyone. Last time, we had got masks, sanitizer and gloves but this time, just nothing.”

“We work as a Sahiya simply because we are in dire need for some money and the small stipend I get by working here, is also a big support for us.”

Says a Sahiya-

“I was compelled to continue my duty despite of myself being infected. At home, my family was worried about my health but I was forced to continue due to fear of losing my job and the stipend. Anyway, we are only given promises and our payments have been never on time. Even to get payments cleared, we still have to pay bribe. Now, I am not supported by my family members to do this work.”

### Information lag

Apart from misinformation, people also hide their infection status. This affected the evolving appropriate control strategies based on local contextual situations.

Says an urban ASHA in this context-

“Many families hide their symptoms of fever and cold, with the fear of being taken away by the health services for having Corona. We did our duties of convincing such families because we do not have the option to leave this job”.

## Discussion and Conclusions: Health workers and Encounter with the Pandemic

Based on the narratives as well as representations, we could identify interlinked dominant and minor specific-context related issues which are important for an equity based universal health coverage. Firstly, training of primary health workers in Primary Emergency Health care is important in order to counter unpleasant human interactions and also for maintaining security. The training is also important to counter misinformation which is hampering positive health actions. Secondly, it is important to reinvigorate the medical loop and preventive protocols in health programs to strengthen the health service system at the grassroots level especially enhance the trust between the workers and the people. COVID 19 pandemic is an opportunity to recognize and reinforce the role of primary care workers and formulate gender sensitive and effective control strategies.

### Training Deficit

We could identify a training and facility deficit in some cases as proper emergency training and handling of PPE was missing. Most of the training was carried out through social media and there was no quality check on such a process. Training is most important as most of the information that the workers gathered are through self-training and internet. This was necessitated owing considerable demand from the people through raising questions and doubts regarding the infection. This is also necessary to counter misinformation which was abundant in many places. Training should also include human management in order to counter unpleasant human interactions. Apart from information, training in Primary Emergency Health Care need to be imparted to the primary level workers and supervisors on a periodic basis.

### Trust Trajectories and Transactions

It is surprising that even in a so-called health and politically conscious society, one could identify a number of instances of violence against primary health workers. It is due to a deficit largely because of the emergence of dominant private sector and alienation of the government health workers. A number of instances highlighted here show the sudden salience of the government sector in pandemic control. It is important to **reinvigorate the medical loop and preventive protocols** in health programs to strengthen the health service system at the village level. Largely, the instances encountered in this case study referred to adhering to specific practices and other emergency measures such as quarantine and undergoing tests. Emergence of misinformation is also due to the trust deficit apart from the dominance of social media.

### Community Protocoling

It is known that health workers could not adequately prepare the community due to lack of information and paucity of time. Given the fact that for many health workers this was the first encounter with a pandemic, it is necessary to evolve a module which gives more focus on programmatic components and system-society interface in order to handle emergency care in future.

## Data Availability

All data pertaining to the study are available for scrutiny

*(The paper was prepared for the Independent Panel for Pandemic Preparedness and Response for presentation in the World Health Assembly)*

